# Effectiveness of control strategies for Coronavirus Disease 2019: a SEIR dynamic modeling study

**DOI:** 10.1101/2020.02.19.20025387

**Authors:** Jinhua Pan, Ye Yao, Zhixi Liu, Mengying Li, Ying Wang, Weizhen Dong, Haidong Kan, Weibing Wang

## Abstract

**Background:** Since its first case’s occurrence in Wuhan, China, the Coronavirus Disease 2019 (COVID-19) has been spreading rapidly to other provinces and neighboring countries. A series of intervention strategies have been implemented, but didn’t stop its spread.

**Methods:** Two mathematical models have been developed to simulate the current epidemic situation in the city of Wuhan and in other parts of China. Special considerations were given to the mobility of people for the estimation and forecast the number of asymptomatic infections, symptomatic infections, and the infections of super-spreading events (I_sse_).

**Findings:** The basic reproductive number (R_0_) was calculated for the period between 18 January 2020 and 16 February 2020: R_0_ declined from 5.75 to 1.69 in Wuhan and from 6.22 to 1.67 in the entire country (not including the Wuhan area). At the same time, Wuhan is estimated to observe a peak in the number of confirmed cases around 6 February 2020. The number of infected individuals in the entire country (not including the Wuhan area) peaked around February 3. The results also show that the peak of new asymptomatic cases per day in Wuhan occurred on February 6, and the peak of new symptomatic infections have occurred on February 3. Concurrently, while the number of confirmed cases nationwide would continue to decline, the number of real-time COVID-19 inpatients in Wuhan has reached a peak of 13,030 on February 14 before it decreases. The model further shows that the COVID-19 cases will gradually wane by the end of April 2020, both in Wuhan and the other parts of China. The number of confirmed cases would reach the single digit on March 27 in Wuhan and March 19 in the entire country. The five cities with top risk index in China with the exclusion of Wuhan are: Huanggang, Xiaogan, Jingzhou, Chongqing, and Xiangyang city.

**Interpretations:** Although the national peak time has been reached, a significant proportion of asymptomatic patients and the infections of super-spreading events (I_sse_) still exist in the population, indicating the potential difficulty for the prevention and control of the disease. As the Return-to-Work tide is approaching and upgrading, further measures (e.g., escalatory quarantine, mask wearing when going out, and sit apart when taking vehicles) will be particularly crucial to stop the COVID-19 in other cities outside of Wuhan.

**What was already known about the topic concerned:** Currently, a Coronavirus Disease 2019 (COVID-19) is thought to have emerged into the human population in Wuhan, and cases have been identified in neighboring provinces and other countries. In existing epidemiological studies, the basic reproduction number (R_0_) of the virus were estimated between 1.4 and 5. Besides, it is of crucial importance to evaluate and improve different intervention strategies which have already implemented.

**What new knowledge the manuscript contributes:** In this study, two mathematical models were established to simulate the current epidemic situation and predict the future trend of the COVID-19. We found that with the implementation of different policies, R_0_ continued to decline over time and the number of confirmed cases in Wuhan will peak on around February 6. Also, we estimated and forecast the number of asymptomatic infections, symptomatic infections, and infections of super-spreading events caused by the COVID-19 and the risk index of different cities.

**Implications of all the available evidence:** Our research has important practical implications for public health policy makers. Although the current prevention and control measures have made some significant inroads into controlling the epidemic, complete control has not yet been achieved. We recommend that self-isolation at home be strictly observed for a period of time in the future. Furthermore, our estimation of the number of asymptomatic people, super spreading and real-time inpatients would provide basis guidance for the hospital to arrange beds accordingly.

## BACKGROUND

On 12 December 2019, a case of pneumonia of unknown etiology was detected in Wuhan City, Hubei Province, China. On 31 December 2019, the outbreak was first reported to the World Health Organization (WHO)(1). The novel coronavirus has thus been named ‘severe acute respiratory syndrome coronavirus 2’ (SARS-CoV-2), while coronavirus disease associated with it is now referred to as Coronavirus Disease 2019 (COVID-19). Most cases from the initial cluster had an epidemiological association with a live animal market (Huanan Seafood Wholesale Market, henceforth referred to as the Seafood Market), suggesting that the SARS-CoV-2 is animal-derived(2). However, the specific source of the virus has not be identified yet.

The reported data show that while it is susceptible to people of all ages, the elderly and those with underlying diseases(3) tend to become more seriously ill once infected(4). The outbreak of COVID-19 started at the time when hundreds of millions of Chinese people were on their way to their hometowns or vacation destinations for the celebration of Chinese New Year, which aided the rapid spread of the virus. As a result, a first-level response to major public health emergencies was launched in many areas across the country. Wuhan, as the source of this outbreak, implemented travel ban on 23 January 2020(5). The Chinese government issued extension of order to shut down all non-essential companies, including manufacturing plants, in Hubei Province until at least 20 February. As of 25 February, 78,190 cases of COVID-19 (according to the applied case definition) have been reported, with 2,718 deaths reported(6).

Since the epidemic outbreak, Wuhan has built two large hospitals with 2,600 beds to ease the sudden increase of local peoples’ healthcare needs. Interventions have also been adjusted for treating milder infections, whereby individuals have been shifted from home-based quarantines to centralized quarantines at designated facilities (e.g. “square cabin” hospitals) in fixed sites, to efficiently monitor and manage infections and prevent further outbreaks. As an intervention strategy, quarantine has been implemented nationwide to prevent further city-to-city transmission. However, despite these measures, the number of daily reported confirmed new cases is still very big.

Most people in China and those who may affected by the COVID-19 outbreak around the world are eager to know when they can return to normal lives. The purpose of this study is to provide a scientific projection on COVID-19 progression. The mathematical models could explain the epidemiological patterns of infectious diseases and provide valuable information for the formulation and evaluation of decision-making in disease preventive(7). This study also estimated the basic and effective reproduction number of the COVID-19, as well as the peak time and size of the epidemic with a dynamic model, and simulate the impact of various intervention strategies, including travel restrictions and voluntary quarantine.

## METHODS

### Data source

#### Reported data

Number of confirmed cases per day from 16^th^ January 2020 to 16^th^ February 2020(8, 9) reported by the National Health Commission of the People’s Republic of China (Only cases that tested positive for nucleic acid were included), the Provincial Health Commissions of China, and government reports from Hong Kong, Macao, Taiwan and other countries. Our data also include the daily numbers of cured cases and deaths. The demographic data is from the Statistical Yearbook of the National Bureau of Statistics and from the Provincial Bureau of Statistics.

### The Models

According to the current epidemic situation and the intervention strategies, the following SEIR mathematical models were constructed. **The Wuhan Model** was based on the epidemic situation in Wuhan from 16 January 2020 to 16 February 2020 and reflected the changes of the infections in the city. It included seven population groups for the seven stages in the progression of the COVID-19. According to the characteristics of the disease, when a susceptible person (Susceptible, S) effectively encounters a person that is infected with the COVID-19, S would be infected with the virus (Exposed, E) and would progress to infectious stage. The infected individual might exhibit symptomatic infection (I) or asymptomatic infection (U), and might cause an infection of super-spreading event (I_sse_); they might get medical care (Treatment, T), and then mostly were shifted to the recovery group (Recovery, R). Notably, the exposed individuals were infected, and had the ability to infect other susceptible individuals. The I_sse_ means those infections can cause three or more generations of infection. The flow diagram for this model is shown in Suppl. Fig 1. **The National Model (except for Wuhan region)** was based on the Wuhan Model, aiming to predict the changes of the Coronavirus infection in other parts of China; the flow diagram for this model is shown in Suppl. Fig 2.

Each model was based on three hypotheses: i) people who had visited or had a contact history with the Seafood Wholesale Market were quarantined, and they did not travel to other parts of the country; ii) I, E and U had the same ability to infect others, while those causing I_sse_ had a greater ability to do so; iii) the daily confirmed cases was diagnosed in hospitals.

### Estimation of epidemiological parameters

In the models, the detection rate was a proportion of confirmed cases from those who were infected. In Wuhan model, we assumed that the detection rate from 16 to 25 January was 10%, the detection rate has been improving since 25 January, with a fixed increment each day. Until the detection rate risen to 75%, the detection rate would not change with time any more. And the detection rate in National model (except Wuhan) were from 0% to 75%, The same as quarantine rate in Wuhan (from 10% to 85%) and National model (except Wuhan) (from 5% to 80%)

In the National Model, parameter a, b were the initial proportion of I group and I_sse_ group, respectively. We used parameter a, b to recalculate the values of μ_1_ (the proportion of U group), μ_2_ (the proportion of I group), μ_3_ (the proportion of I_sse_ group). And finally fitted results showed that the proportion of asymptomatic patients, μ_2_, in the population was 0.0828*a /(0.0828*a+b+a). The same as Wuhan model, we used the least square estimate to estimate the μ_1_ (the proportion of U group). In the National Model, Wuhan data was used to calculate the p of various periods (p stands for the proportion of the latent population in the outflow population of Wuhan).

In the National Model, we assumed that the quarantine rate was 5% on January 16, with a fixed increment every day, and that this rate did not change with time once it reached 80%, after 20 days. We used public data to calculate the proportions of the outflow and inflow population from January 16 to January 23(10), then allocated the number of daily outflows in Wuhan, based on this proportion. All parameters used in the model are shown in ***Table 1***.

According to the incubation proportion p of the daily outflow population from January 16 to January 23 estimated by the Wuhan model and the proportion of daily migration population from Wuhan to other cities, we calculated the cumulative number of people entering each city in the incubation period within 8 days. The risk index is calculated by the product of the cumulative number of people in the incubation period (until February 10, 2020) and the resident population of each city.

### Basic reproductive number (R_0_)

The basic reproduction number R_0_ represents the expected number of secondary cases produced by an initial infectious individual, in a completely susceptible population. The calculation for determining R_0_ was done by using the R software(11).

### Data Availability

The data in this article are all public data obtained from the National Health Commission of the People’s Republic of China.

### Code availability

The code is available from the corresponding author upon reasonable request.

## RESULTS

### Simulation of the Wuhan Model

Diagnosis data (open source) from Wuhan for the COVID-19 from January 16, 2020 to February 16, 2020 were used for the modeling. Figure 1 shows that the Wuhan Model fits well the trend of the number of recent confirmed cases in Wuhan: it predicts that the peak of confirmed cases in Wuhan will be 1,718, and would occur around February 6. The peak of the number of asymptomatic infections appears on February 6, while the number of I_sse_ and symptomatic infections would reach their maximal numbers on February 3, with peak daily number for the three types of infections being 1,017, 419 and 12,418, respectively (Figure 2). And the number of confirmed cases would reach the single digit on March 27 in Wuhan.

**Figure 1.**
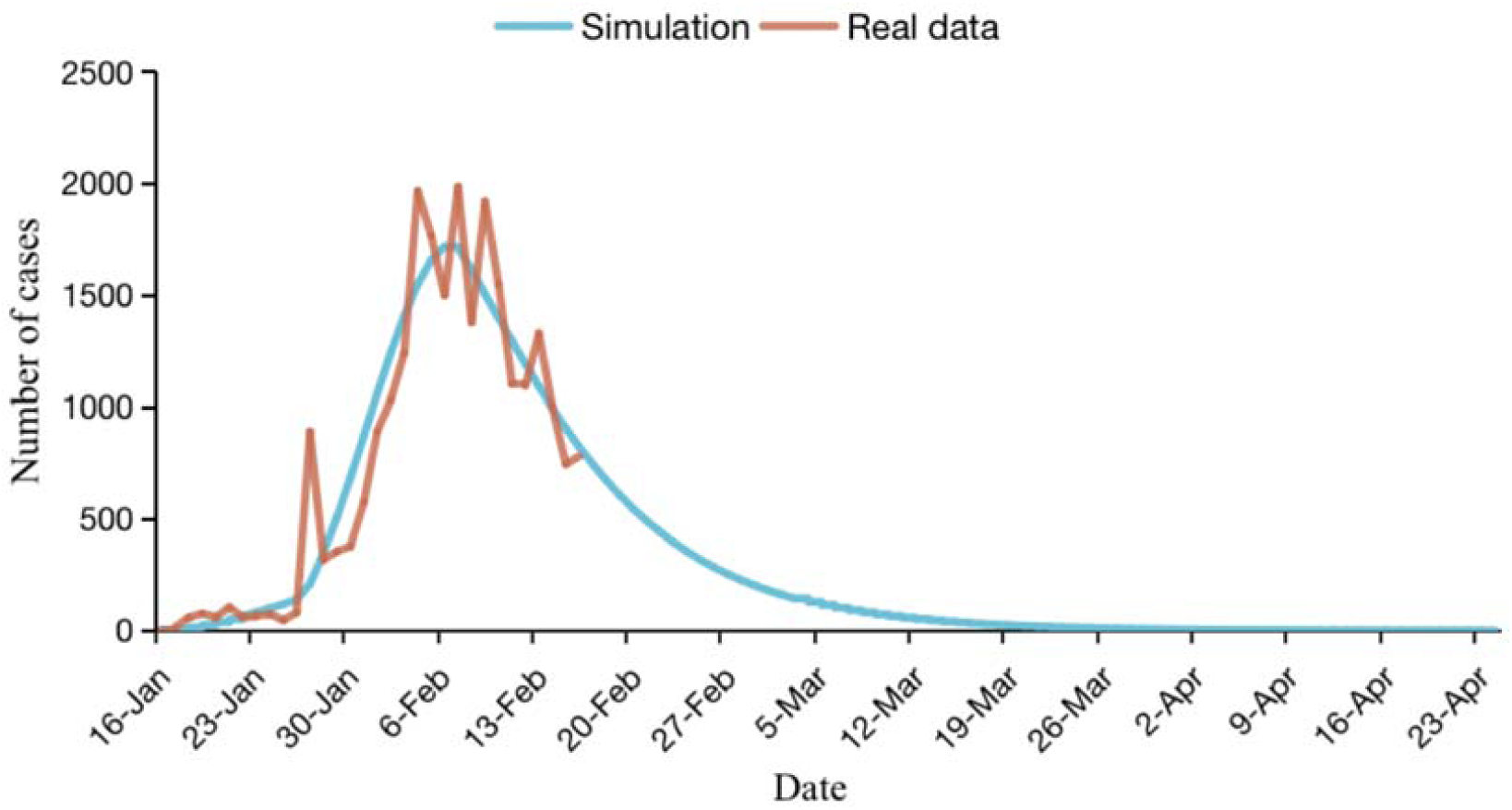
Prediction of the number of new confirmed cases per day. The brown line shows the actual number of newly confirmed cases every day in Wuhan until February 16; the blue line shows the results of model fitting.

**Figure 2.**
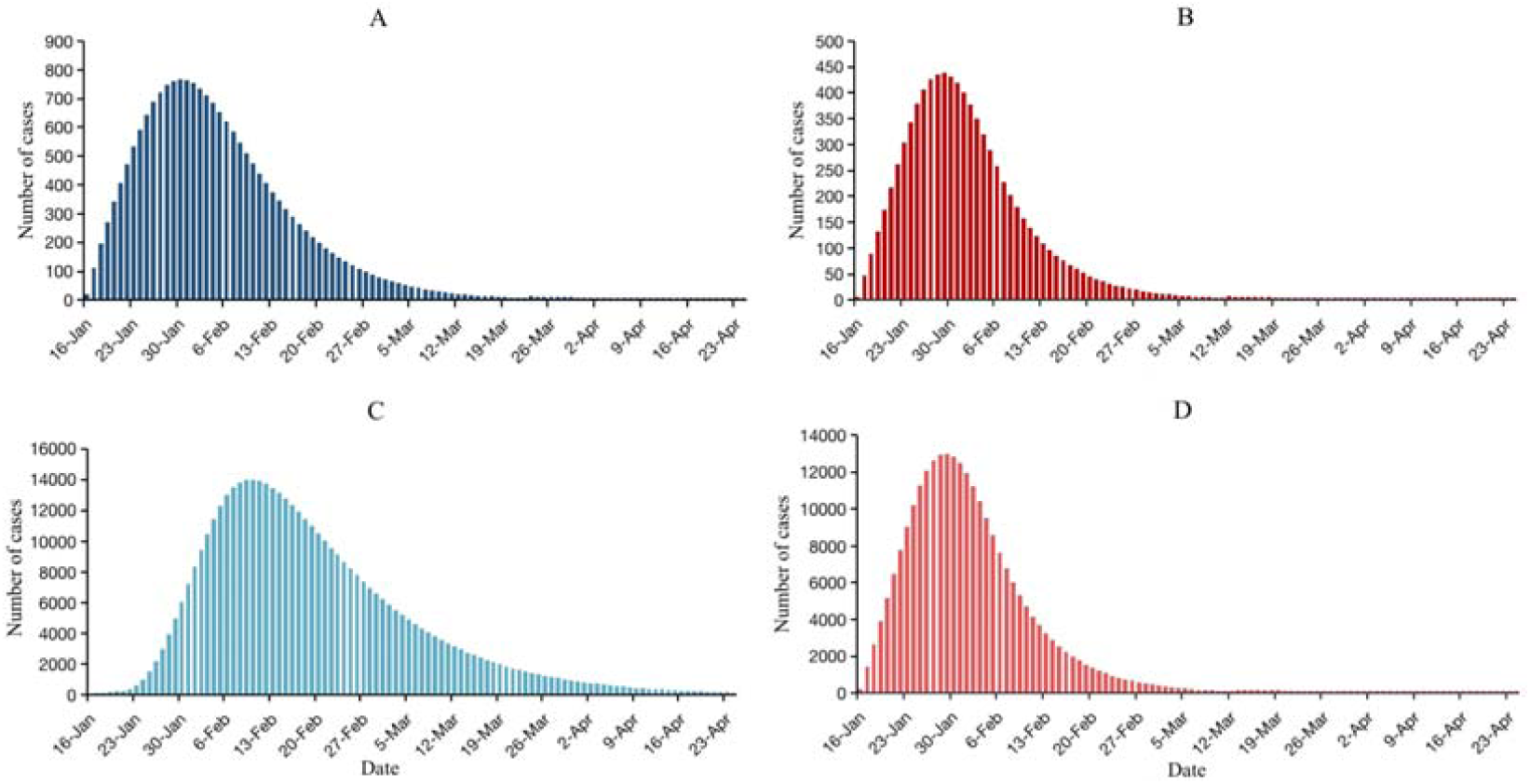
Estimation and prediction of asymptomatic infections, symptomatic infections and super spreading event infections in Wuhan city. **A**: Estimation and prediction of daily asymptomatic infections in Wuhan. **B**: Estimation and prediction of daily super-spreading event in Wuhan. C: Estimation and prediction of the real time inpatients. **D**: Estimation and prediction of the occurrence of new infections per day. The time scale predicted by the model is from 16 February to 24 April 2020.

We established models for various scenarios, as shown in Suppl. Figure 3. The smaller the outflow of the population, the greater the daily number of new confirmed cases and the real time inpatients in Wuhan. Thus, if the outflow of the population is 6 million, the daily number of newly confirmed cases in Wuhan would reach 860. Similarly, a dynamic model was applied to analyze the daily increase in the number of newly confirmed cases in Wuhan. Suppl. Figure 4 shows that the higher the early detection rate, the greater the number of confirmed cases, and the earlier the peak appears.

When the outflow population is kept constant and the quarantine rate increases gradually, the number of newly confirmed cases decreases. To predict the impact of various quarantine rates, we set isolate rate=10% as the baseline number of cases; when setting isolate rate=20%, it reduced to 1,159,720 inpatients cases on the day of peak numbers, a decrease of 11.18%; isolate rate=30% would reduce the number of inpatients cases to 933,070 (Figure 3).

**Figure 3.**
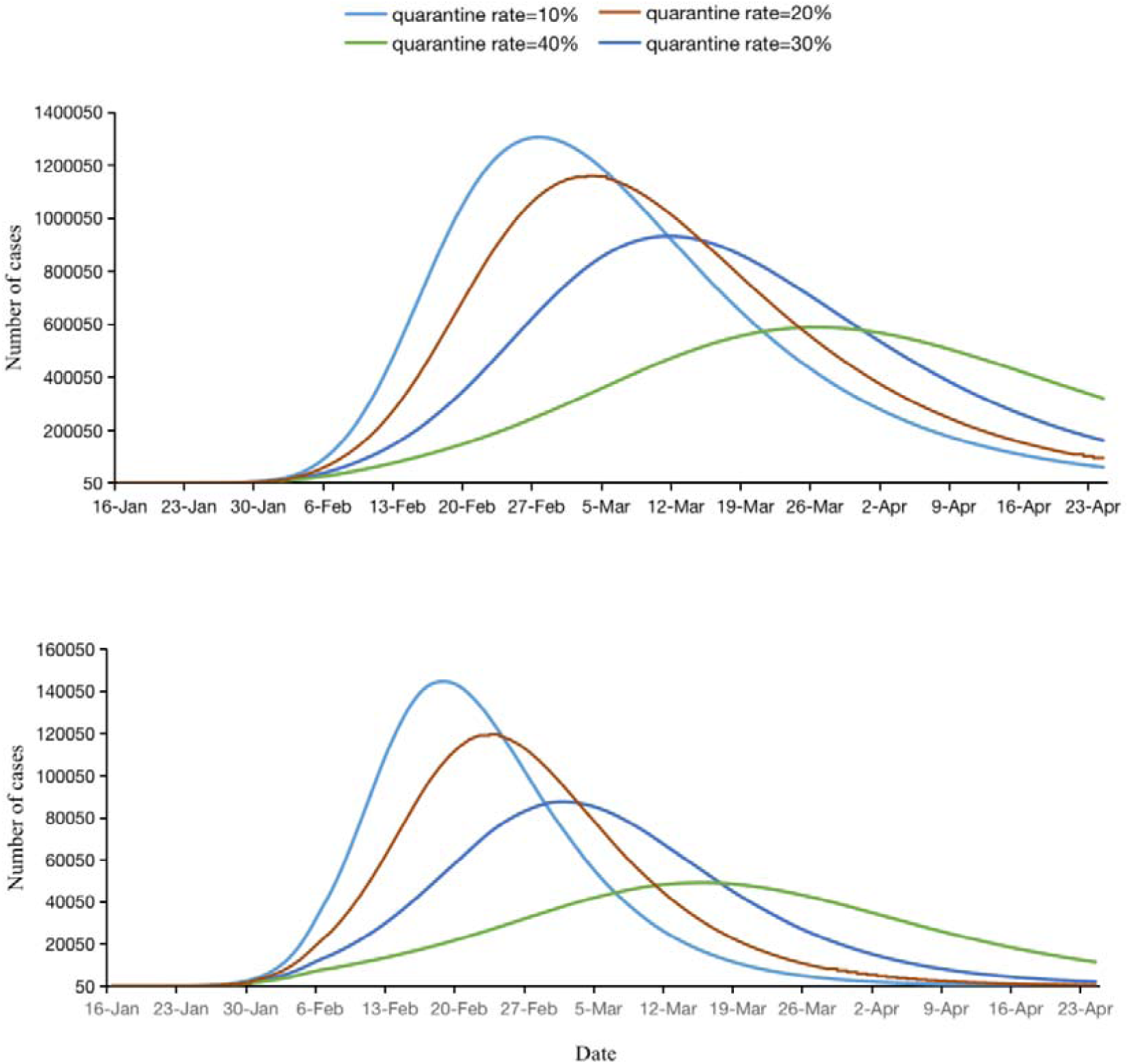
Epidemic forecasts for Wuhan under different scenarios of quarantine rate. **A** and **B** predict the impact of different various quarantine rates on the real time inpatients and the number of newly confirmed cases per day.

In the Wuhan Model, we set a function related to the parameter m and time t as follows: before January 23, 2020, m infected people contacted the Seafood Wholesale Market, while the zoonotic infective cases no longer occurred thereafter due to the closure of that Market. The Model simulated various scenarios to assess the impact on the epidemic situation of an increase in the force of infection by 1, 1.5, 2, 2.5, and 3 times. By predicting the impact of the number of zoonotic infective cases, we set 43 as the number of baseline cases. Doubling the number m (to 86) increased the number of cases by 1408 on the peak day, and enhanced the confirmed cases by approximately 9.10% on that day, while augmenting m to triple the baseline increased the confirmed cases by approximately 22.58% (Suppl. Figure 5).

### Simulation of the National Model (except Wuhan)

Figure 4 shows the fitting of the nationwide mathematical modeling. Our Model rationally fitted the trend of the daily numbers of newly confirmed cases in other parts of China, and predicted that the peak number of confirmed cases would occur around February 3, while the highest number of confirmed cases would reach 1,756 per day. The peak arrival time of the simulation data was consistent with that of the real data.

**Figure 4.**
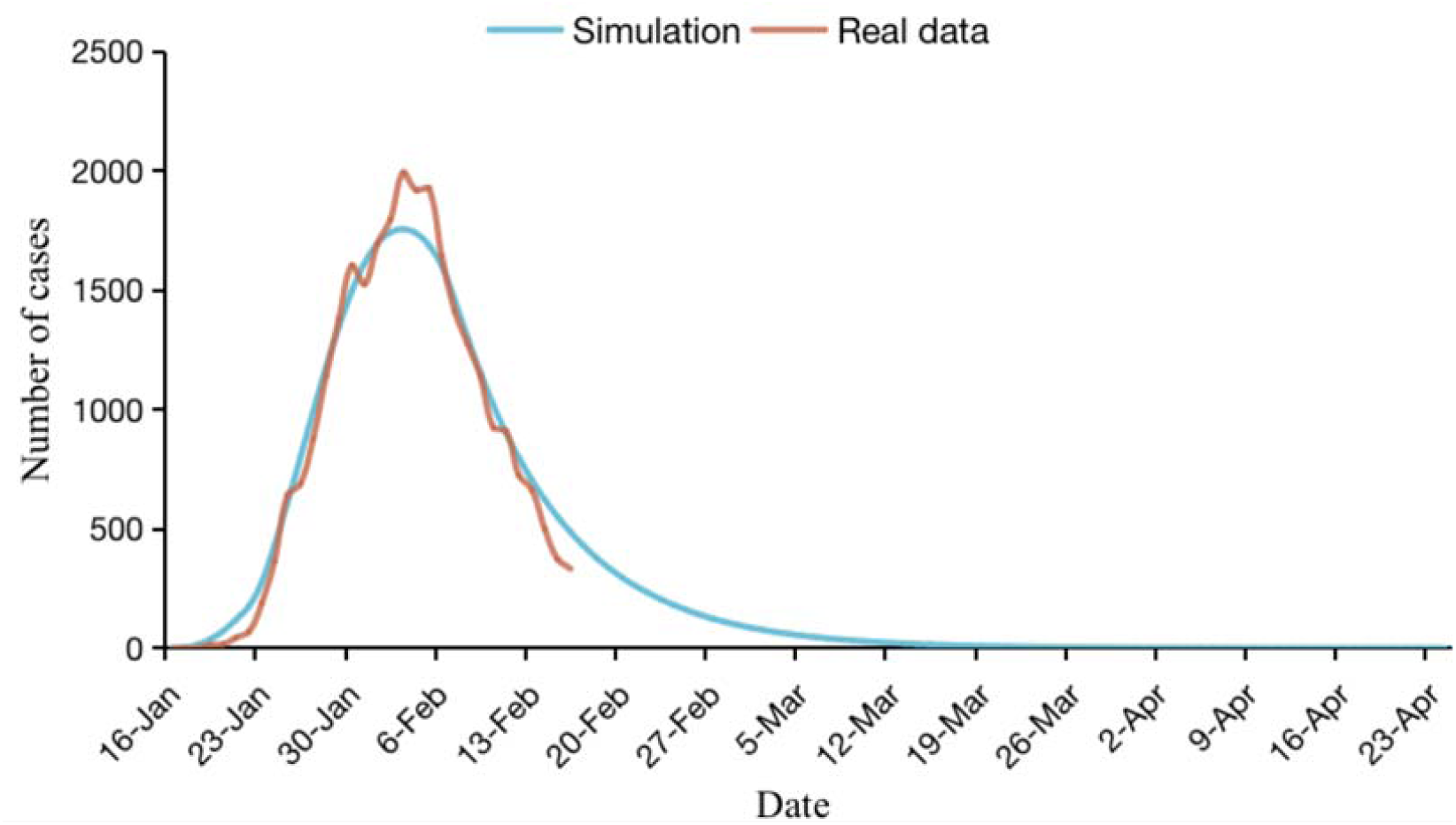
Estimation and prediction of newly confirmed cases in China (except Wuhan). The brown line shows the actual number of newly confirmed cases each day in the cities in China (except Wuhan). The blue line shows the result of model fitting, with the model applied to predict the number until 24 April 2020.

Figure 5 shows that the peaks of asymptomatic infections and I_sse_, and the real time inpatients and symptomatic infections would appear on January 30 and January 29, February 10 and January 29, respectively, with peak number 766, 438, 14,008 and 13,006, respectively. The COVID-19 cases would gradually disappear by the end of April. The number of confirmed cases would reach the single digit on March 19 in the entire country (not including the Wuhan area).

Suppl. Figure 6 shows the positive correlation between population inflow and case numbers per day. When the population inflow is 6 million, the number of new cases per day reaches a peak of 1,838, while inflow become 3 million, the number would be 1,539. An increase of 16.2% over the peak would occur when the population inflow was 6 million. We also found that the change in the inflow population was relatively insensitive to the population flow.

**Figure 5.**
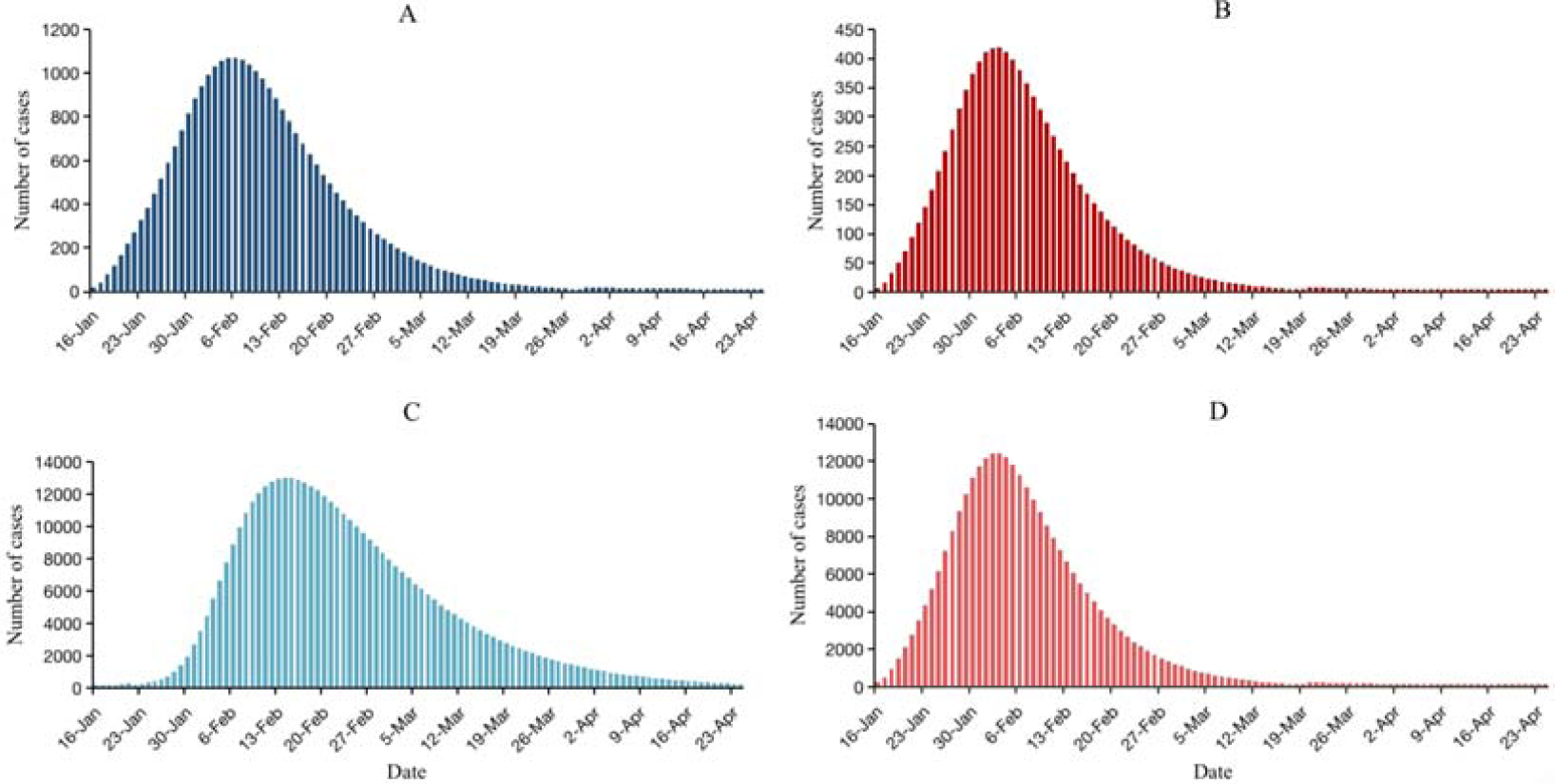
Estimation and prediction of asymptomatic infections, super-spreading event infections, total hospital cases and symptomatic infections in China (except Wuhan). **A:** Estimation and prediction of new asymptomatic infections per day in China (except Wuhan). **B:** Estimation and prediction of new super-spreading event infections per day in China (except Wuhan). **C:** Estimation and prediction of the real time inpatients. **D**: Estimation and prediction of new infections per day among people in China (except Wuhan).

By predicting the impact of various quarantine rates on the number of cases each day, we observed that the peak decreased gradually, and the number of new and cumulative confirmed cases per day had a clear downward trend with the gradual increase of the quarantine rate; when the inflow population remained constant, every 5% increase in quarantine rate reduced the numbers of the cumulative confirmed cases, on average, by 313 cases on the peak day. Furthermore, setting the quarantine rate at 45% would reduce the number by 939 cases on the peak day compared to when the quarantine rate was set at 30% (Figure 6). When the detection rate gradually increased, the peak gradually moved forward, and the number of newly confirmed patients showed a significant downward trend. Also, that every 0.2 increase in the detection rate leads to an advance of 1-2 days at the peak (Suppl. Figure 7).

**Figure 6.**
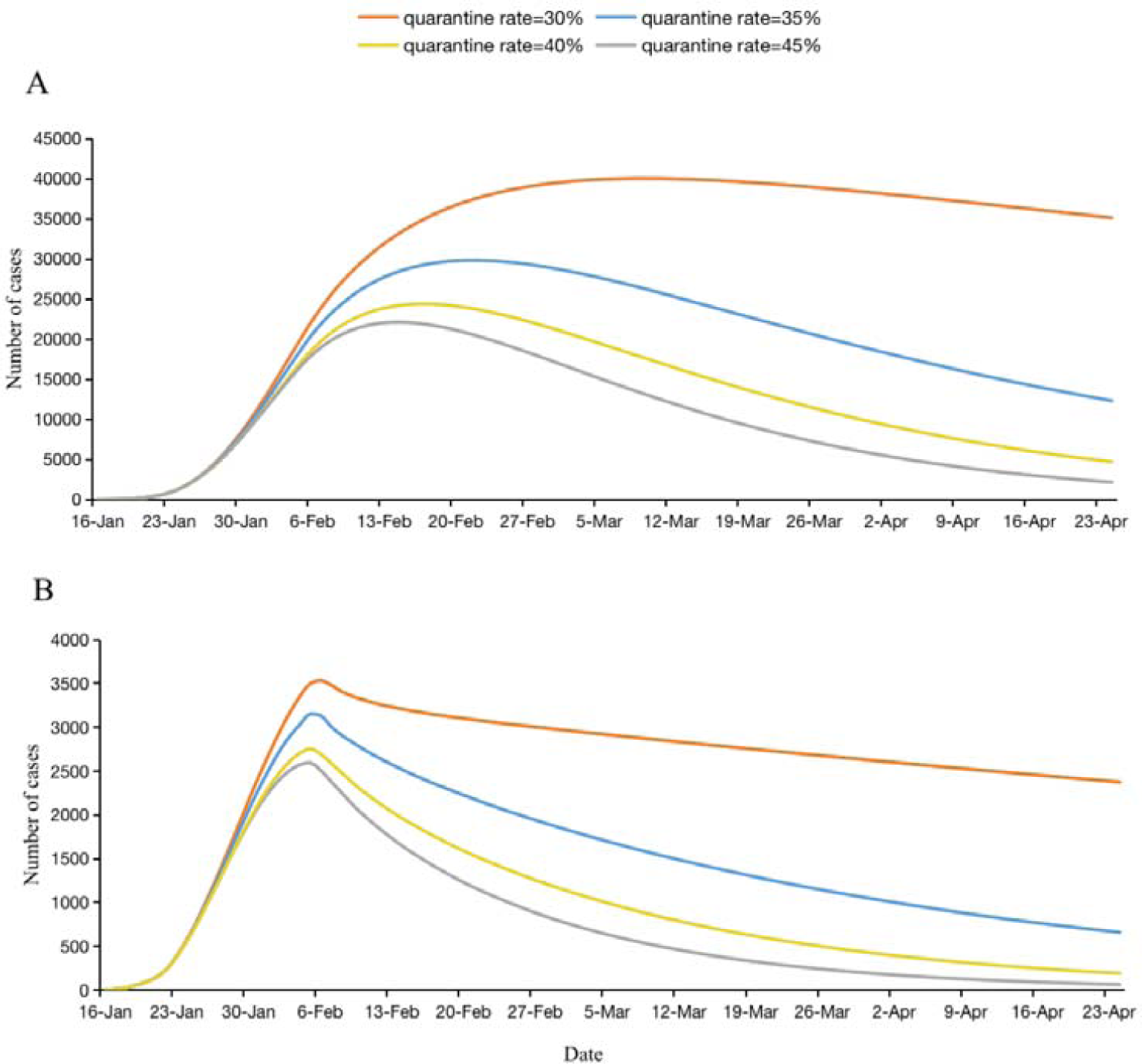
Epidemic forecasts for China (except Wuhan) under different scenarios of quarantine rate. **A** and **B** are used to predict the impact of different quarantine rates on real time inpatients and new confirmed cases per day using national (except Wuhan) models.

Figure 7 shows that the risk index of COVID-19 risk spread by the outflow population of Wuhan to different cities. The top 5 cities ranked by the risk index are Huanggang, Xiaogan, Jingzhou, Chongqing and Xianyang. And The number of incubators from Wuhan to these cities on the 16th to the 23^rd^, 2020 were 474, 513, 249, and 40, 145 cases, respectively. The risk index of 15 cities was consistent with the actual cumulative number of confirmed cases (*r=*0.88, *p*= 3.8*10^^-8^, and the actual cumulative number of confirmed cases were calculated until February 10, 2020).

We have estimated the basic reproduction number R_0_ from 19 January to 16 February 2020 in Wuhan and the whole country (except Wuhan). Figure10 showed that R_0_ declined with time as well as with implementation of different policies, from 5.75 to on 19 January to 1.69 on 16 February in Wuhan, and from 6.22 to 1.67 in the entire country (not including the Wuhan area)

**Figure 7.**
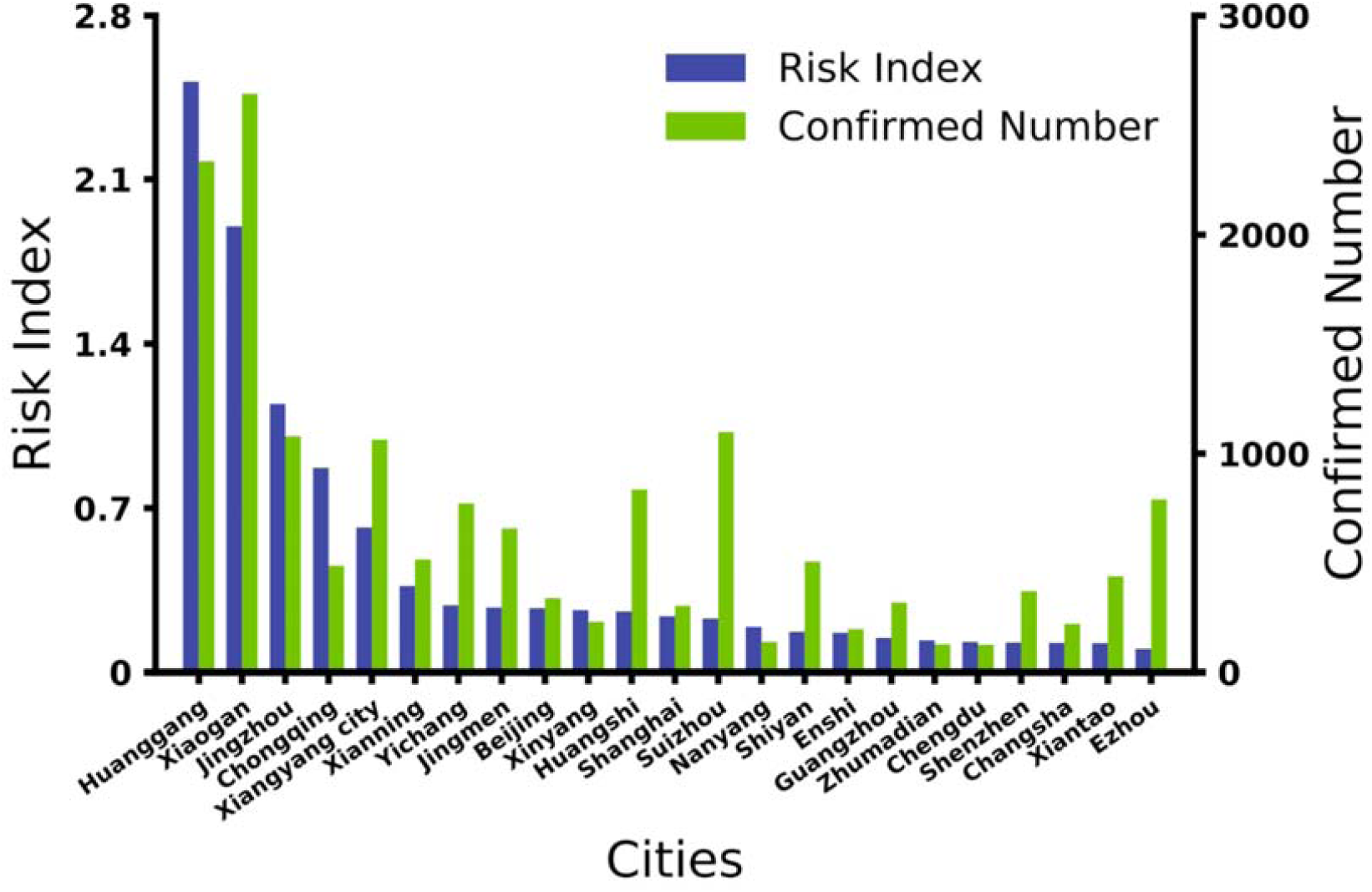
Comparison of the risk index and cumulative number of confirmed cases. According to the incubation proportion p of the daily outflow population from January 16 to January 23 estimated by the Wuhan model and the proportion of daily migration population from Wuhan to other cities, we calculated the cumulative number of people entering each city in the incubation period within 8 days. The risk index is calculated by the product of the cumulative number of people in the incubation period and the resident population of each city. And we compare the risk index with the cumulative number of confirmed cases (*r*=0.88, *p*= 3.8*10^^-8,^ and the actual cumulative number of confirmed cases are calculated until February 10, 2020).

**Figure 8.**
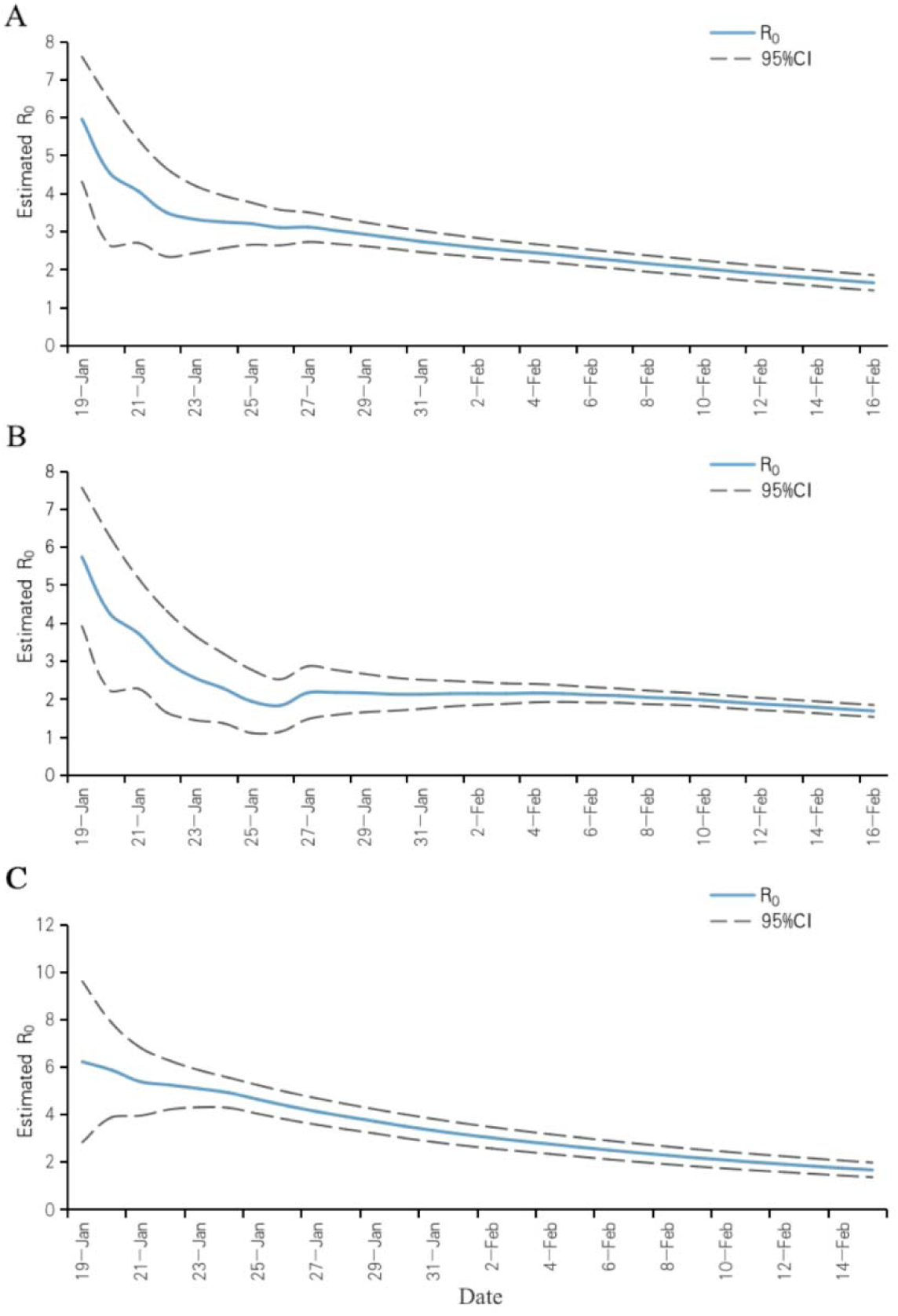
The change of basic reproduction number R_0_. The blue line in the figure represents the estimated value of R_0_ and ts change with time, and the gray line represents the 95% confidence interval of R_0_. **A:** R_0_ and its change with time in China. **B:** R_0_ and its change with time in Wuhan. **C:** R_0_ and its change with time in the entire country except Wuhan

## DISCUSSION

Based on the COVID-19 cases data from 16 January 2020 to 16 February 2020, the data showed that R_0_ declined with time as well as with implementation of different policies, from 5.75 to 1.69 in Wuhan and from 6.22 to 1.67 in the entire country (except Wuhan), which was consistent with other studies(12, 13) but we first estimate the trends. Although R_0_ for the COVID-19 was declined on February 16 in China, it was still higher in China (1.66; 95% CI 1.86–1.46), compared to the range of the R_0_ for SARS epidemic(12, 14), and for MERS(12, 14). On the other hand, such a high number for R_0_ was consistent with the opinion that the virus had gone through at least three-to-four generations of transmission and suggested that there might be a possibility of aerosol transmission as SARS did(15), though whether there was aerosol transmission needs further investigation.

Nowcasting and forecasting are of crucial importance for public health planning and control, both domestically and internationally. Our models evaluated the epidemic situation in Wuhan and in other parts of China separately, with the consideration of various intervention measures. The results of our modeling suggest that Wuhan have reached a peak in the number of confirmed cases around 6 February, with the maximum number of confirmed cases at 1,718 per day, while the peak of the number of confirmed individuals in the whole country has already occurred around February 3 and will reach single digit on March 27 in Wuhan and March 19 in in the whole country except Wuhan. In addition, our study predicts that the number of inpatients in Wuhan reached a peak of 13,030 on 14 February, and begin to decline, which can provide certain basis for hospital bed arrangement.

Model fitting for the newly confirmed cases per day gave a number that was lower than the actual number, and the peak number of newly confirmed infected cases per day in the model fitting of the entire country except Wuhan was also lower than the actual number – these discrepancies may be due to underreporting, and delayed diagnosis during the early stages of the epidemic. The underreporting and insufficient diagnosis might be due to less nucleic acid test kits, false negative result of the testing(16), and asymptomatic patients(3). Due to the recent change of more availability of test kits and greater efforts of various government departments, the detection capacity has continuously improved, leading to a sharp rise in patients’ numbers compared to the models’ estimates.

Current interventions, including extended vacations and self-quarantine at home, aim to isolate high-risk populations. We have analyzed the different quarantine rates, to determine how this variation changes the epidemic trend. Assuming that the major cities in the country have not taken the first level response, the isolation rate is not as high as it is currently, we reduced the quarantine rate and simulate the situation that if the country has not taken the first level response in Wuhan. When the quarantine rate was 10%, the peak value of newly confirmed cases would be 1,305,713 in Wuhan; when the quarantine rates became 20%, 30% and 40%, the peak value of newly confirmed cases would reach 1,159,720, 933,070 and 588,814 in Wuhan respectively. The scenario suggested that Wuhan ‘s mandatory isolation and first-level response had played a great role in the stop of epidemic spreading. Many cities in the country had started the first level response. We reduce the isolation rate in the National model to the scenario that the first level response was not launched. When the quarantine rates are 30%, 35%, 40%, 45%, the peak value of newly confirmed cases will reach 3,532, 3,151, 2,753, and 2,593 in the entire country. And if the quarantine rate is less than 30%, the new confirmed cases will grow explosively. Comparing to the current situation with a maximal quarantine rate of 80% in the National Model, it adds up 837 confirmed cases each day at least, which shows the quarantine intervention and first level response measures taken by some cities in China are indispensable policies and show positive effect on the control of the COVID-19. The Return-to-Work tide is approaching, especially in major cities like Shanghai and Shenzhen, upgraded measures (*e*.*g*., escalatory quarantine, mask wearing when going out, and sit apart when taking vehicles) will be particularly crucial for the blockage of the COVID-19. We assumed that the isolation rate in Wuhan will reach 85% and 80% in the whole country except Wuhan. However, if we further improve the isolation rate, the epidemic will end ahead of time.

A proportion of the first generation of infected people had visited the Seafood Market in Wuhan(4). Due to the existence of asymptomatic infections and the possibility of missed diagnoses, rather than analyzing just the number of people who have been to the Seafood Market, we have instead analyzed the number of people who have contact history with the Seafood Market; when this alternative situation is taken into consideration, the greater the number of people who visited the Seafood Market in the early stage the bigger is the number of people infected, with a higher peak value. However, this relationship is not sensitive. While people at the Market were found and isolated and were no longer contagious, the scale of transmission of this first generation of virus might be larger than that of other infected individuals.

This study primarily focused on the situation that can be judged from the appearance of some infection symptoms, such as fever, dry cough, etc(17). According to the latest research results of Dr. Zhong Nanshan, they proved the presence of another type of infected people – those who are asymptomatic infections at COVID-19 diagnosis(3). According to the results of the Wuhan Model and National model(excluded Wuhan), there are indeed asymptomatic cases with peak values 1,071 in Wuhan (766 in the other parts of China) and I_sse_ cases with peak values 419 cases in Wuhan (438 in the other parts of China), asymptomatic cases are hidden virus carrier, finding them is extremely difficult but this population will increase in number rapidly, which is particularly challenging in terms of prevention and control. The only effective intervention for asymptomatically infected people is to prevent close personal contacts. Concurrently, the number of real-time COVID-19 inpatients in Wuhan has reached a peak of 13,030 on February 14 before it decreases (14,008 on February 10 in the other parts of China), which can provide some basis for the arrangement of hospital beds. The model further shows that the COVID-19 cases will gradually disappear by the end of April 2020, both in Wuhan and the other parts of China. The number of confirmed cases would decrease to less than 10 on March 27 in Wuhan and March 19 in the other parts of China.

During the current epidemic outbreak, the public should be reminded not only to practice at-home quarantine, but also to pay attention to personal protection, *e*.*g*. wearing masks, and reducing communicating with other people in person, especially after the stage of high level response (travel restricts, the postponement of school semesters, *etc*.). As the potential long incubation period of the COVID-19, the effects of public health measures, such as to place someone and his/her close contacts under medical quarantine at both the hospital and community, would also depend on the swift and accurate diagnosis – any delay will make the public to pay a high cost(18).

Our study has several limitations. First, there is no enough data to provide accurate information about quarantine rates and detection rates at each stage of the epidemic, as the denominator could also be underestimated due to limited medical source for treatment, false positive testing kits. However, we have adjusted for the initial values of the parameters in the models based on the available survey data. Second, based on the changes in prevention and control modes and societal efforts at each stage, we have estimated the functions of the quarantine and detection rates in relation to time, in order to simulate the actual conditions as closely as possible. In addition, we were unable to obtain some relevant data that may be very important to the study, such as the exposure and possible infection rates of healthcare workers nationwide(19). Instead, we use the estimation for exposure and infection rate of that population from a survey (unpublished data) based on a big specialized hospital in Wuhan.

## CONCLUSIONS

China’s prevention and control measures have made significant inroads into controlling the epidemic of COVID-19, but the complete control has not yet to be achieved. This study found self-quarantine at home should be strictly observed in the future, and that the quarantine level to be maintained at a relatively high level to prevent the possibility of a second outbreak of the epidemic. When the accuracy and sensitivity has been improved substantially, *e*.*g*., by using CT scan instead of depending only on testing kits, the continued success may still rely on the community screening in order not to miss any and effectively cut the transmission chain from those with mild symptoms. Furthermore, professional medical care personnel should supervise centralized quarantine at certain quarantine sites, to better monitor and manage infected peoples and prevent further expansion of infection and improve the health of the patients. Given the high rate of infectivity of this new coronavirus, and there is still a possibility of the virus being spread globally, the countries other China also need to mobilize enough medical supplies to deal with this massive outbreak based on local risk assessments.

## Author contributions

J.P., Z.L., Y.Y. and W.W. designed the study. J.P., Z.L., Y.Y. M.L, and Y.W. collected COVID-19 incidence data and gained insight into the biology and natural history of the virus. J.P., Z.L., H.K. and W.W. developed the model and obtained the related parameters. Y.Y. revised and improved the model, W.D. revised the language, J.P., Z.L., and M.L drafted the manuscript. All authors critically reviewed and approved the final version of the manuscript.

## Acknowledgments

We thank Dr. Yang Liu from London School of Hygiene & Tropical Medicine, Faculty of Epidemiology and Population Health, Department of Infectious Disease Epidemiology for modeling support. This study is sponsored by the Bill & Melinda Gates Foundation (OPP1216424) and Shanghai Sailing Program (Grant No. 17YF142600).

## Supplementary information

**Supply. Table 1.** Parameters of Wuhan dynamic model.

| Parameter | Description | Value | Source |
| --- | --- | --- | --- |
| $d$ | Transition rate of latent individuals to infections | 1/7 | Public data |
| $\gamma_1$ | Transition rate from hospitalization to recovery | 1/14 | Public data |
| $\gamma_2$ | Transition rate of asymptotical infections to recovery | 1/7 | Assumed |
| $\beta_1$ | Transmission coefficient of I, U, and E | 0.4098 | Least square method |
| $\beta_2$ | Transmission coefficient of $I_{sse}$ | 1.0245 | Least square method |
| $\beta_3$ | Transmission coefficient of South China Seafood Market | 3.8369 | Least square method |
| $\lambda$ | Transition rate of infectious individuals to quarantined infections | 1/4.95 | Public data |
| $\mu_1$ | The proportion of U compartment | 0.08 | Least square method |
| $\mu_2$ | The proportion of I compartment | 0.89 | Public data |
| $\mu_3$ | The proportion of $I_{sse}$ compartment | 0.03 | Public data |
| Rate_quarantine(t) | Quarantined rate of individuals | Change with time | Assumed |
| Rate_detection(t) | detected rate of infection individuals | Change with time | Assumed |
| Rate_emigration(t) | emigrated rate of susceptible individuals and exposed individuals | Change with time | Assumed |
| $\alpha_1$ | Death rate of COVID-19 cases in Wuhan city per day | 0.0530 | Reported data(8, 9) |

**Supply. Table 2.**
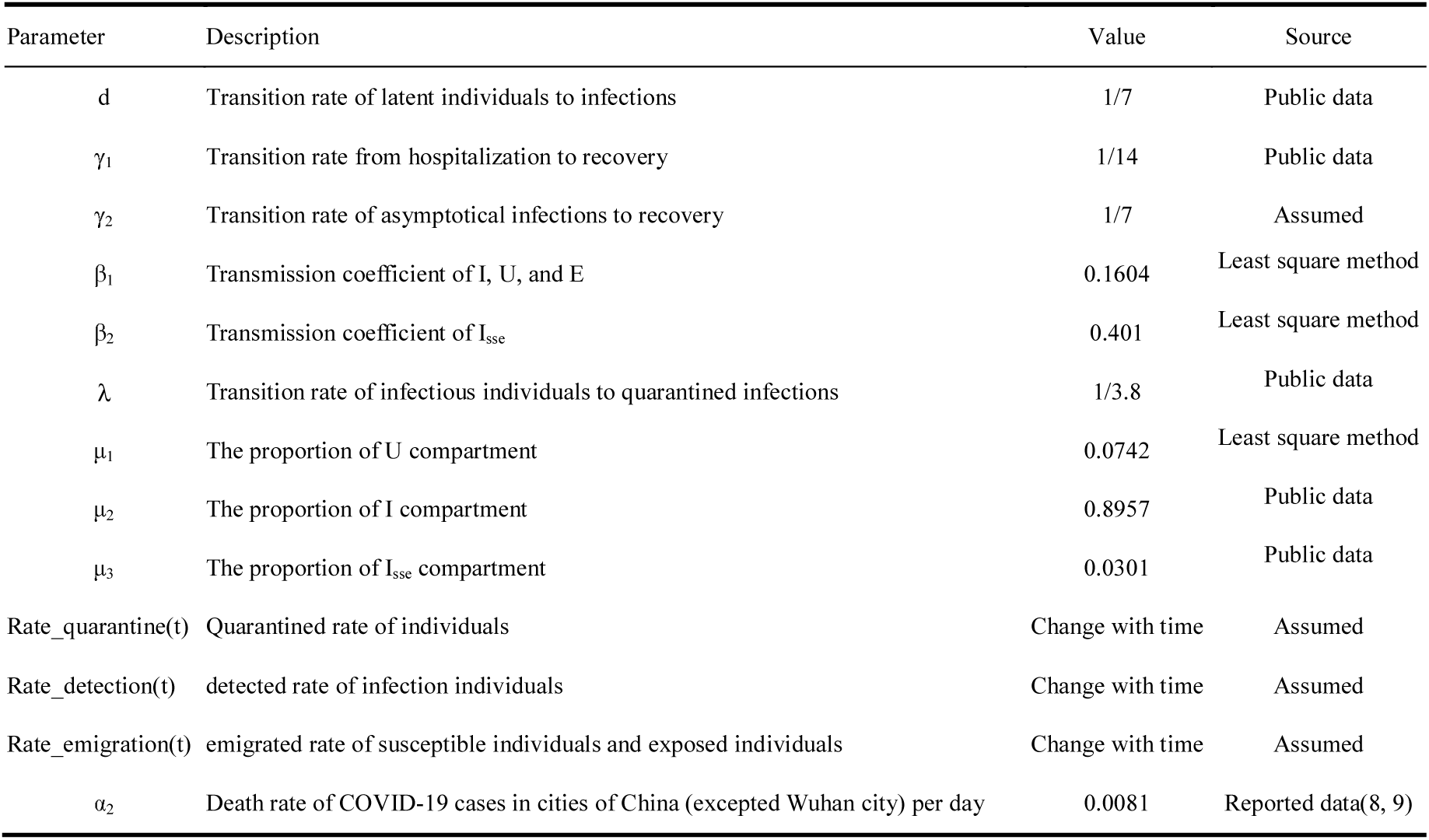
Parameters of National dynamic model except Wuhan.

| Parameter | Description | Value | Source |
| --- | --- | --- | --- |
| $d$ | Transition rate of latent individuals to infections | 1/7 | Public data |
| $\gamma_1$ | Transition rate from hospitalization to recovery | 1/14 | Public data |
| $\gamma_2$ | Transition rate of asymptotical infections to recovery | 1/7 | Assumed |
| $\beta_1$ | Transmission coefficient of I, U, and E | 0.1604 | Least square method |
| $\beta_2$ | Transmission coefficient of $I_{sse}$ | 0.401 | Least square method |
| $\lambda$ | Transition rate of infectious individuals to quarantined infections | 1/3.8 | Public data |
| $\mu_1$ | The proportion of U compartment | 0.0742 | Least square method |
| $\mu_2$ | The proportion of I compartment | 0.8957 | Public data |
| $\mu_3$ | The proportion of $I_{sse}$ compartment | 0.0301 | Public data |
| Rate_quarantine(t) | Quarantined rate of individuals | Change with time | Assumed |
| Rate_detection(t) | detected rate of infection individuals | Change with time | Assumed |
| Rate_emigration(t) | emigrated rate of susceptible individuals and exposed individuals | Change with time | Assumed |
| $\alpha_2$ | Death rate of COVID-19 cases in cities of China (excepted Wuhan city) per day | 0.0081 | Reported data(8, 9) |

**Suppl. Figure 1.**
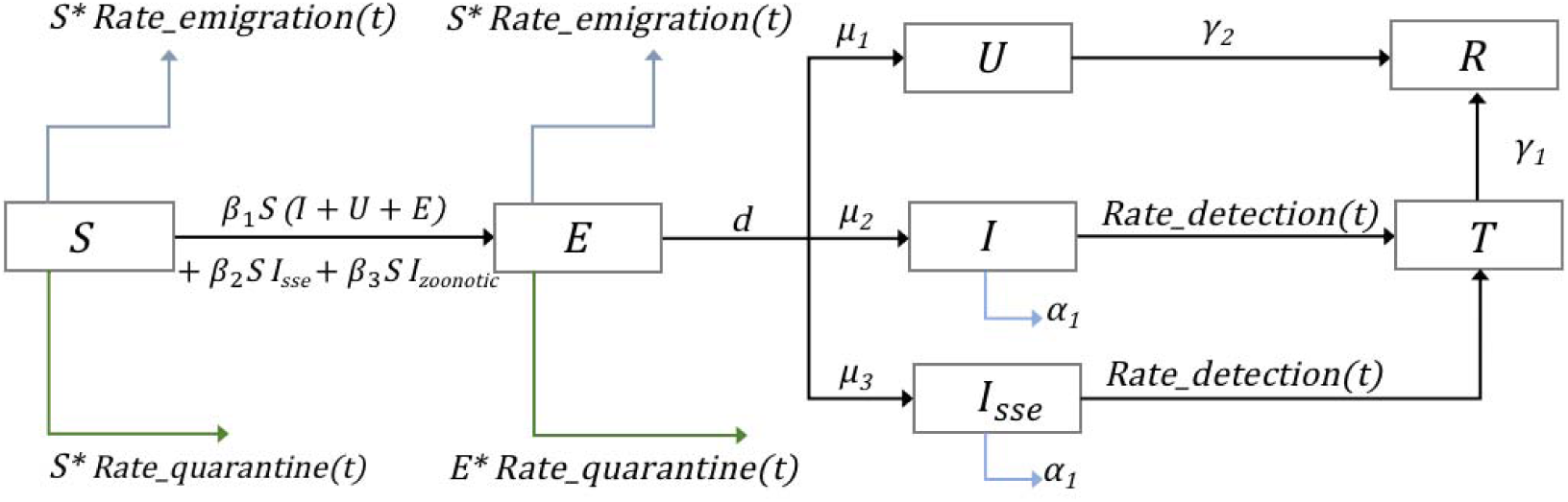
Flow diagram of the Coronavirus Disease 2019 model for Wuhan.

**Suppl. Figure 2.**
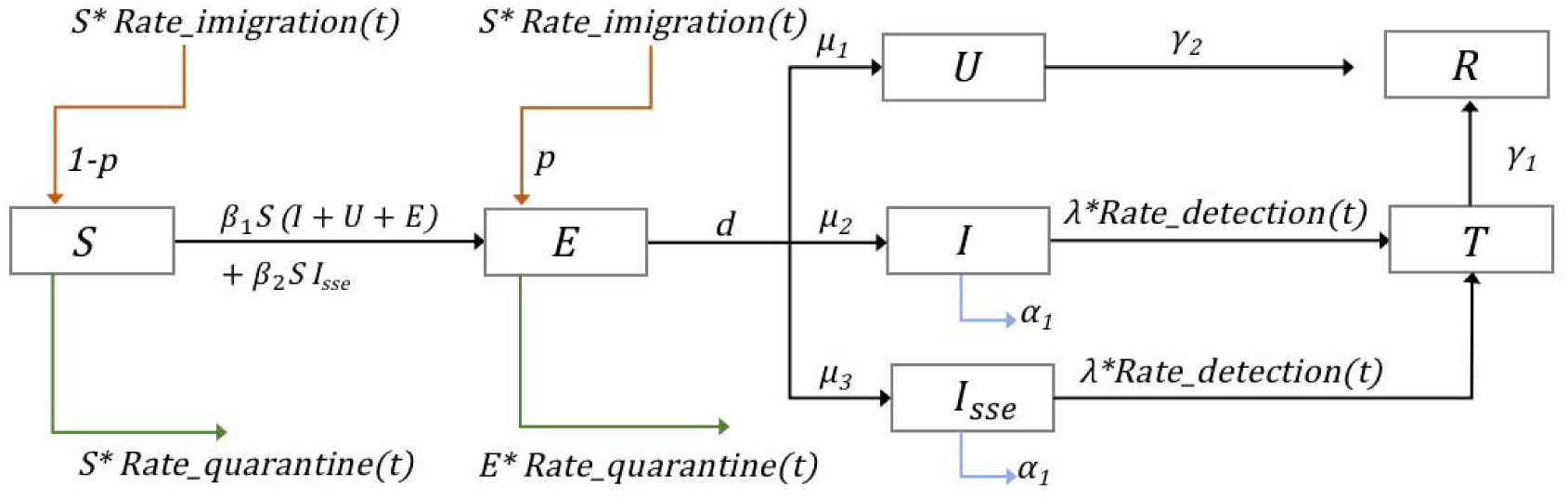
Flow diagram of the Coronavirus Disease 2019 model for the Nation (except Wuhan)

**Suppl. Figure 3.**
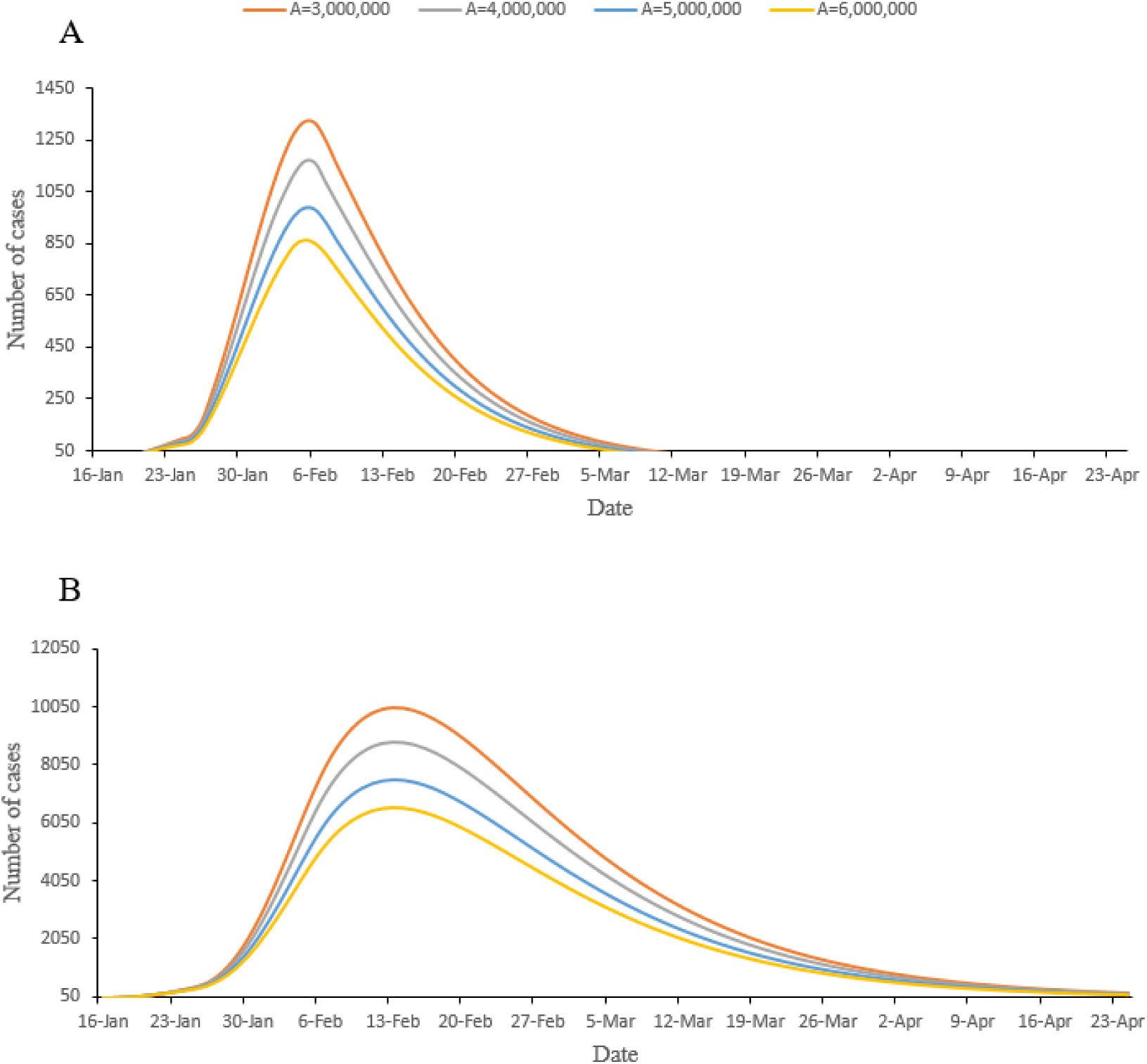
Prediction of new conformed cases and total cases diagnosed in hospital per day in different A values. The dynamic model for analyzing different situations of A, where A represents the total outflow population of Wuhan from 16 January to 23 January. We used public data to calculate the proportion of the outflow population (outflow population rate per day are 9%,9%,10%,10%,12%,15%,18%,17%) from 16 January to 23 January 2020 in Wuhan, then allocated the number of daily outflows in Wuhan, based on this proportion. For this figure, we assumed the number of daily outflows at different levels and observed the number of new confirmed cases as well as the cumulative number of confirmed cases of the 2019 novel coronavirus. Blue, orange, gray, and yellow lines represent a total of 3,000,000, 4,000,000, 5,000,000 and 6,000,000 people, respectively, having flowed out of Wuhan in 8 days.

**Suppl. Figure 4.**
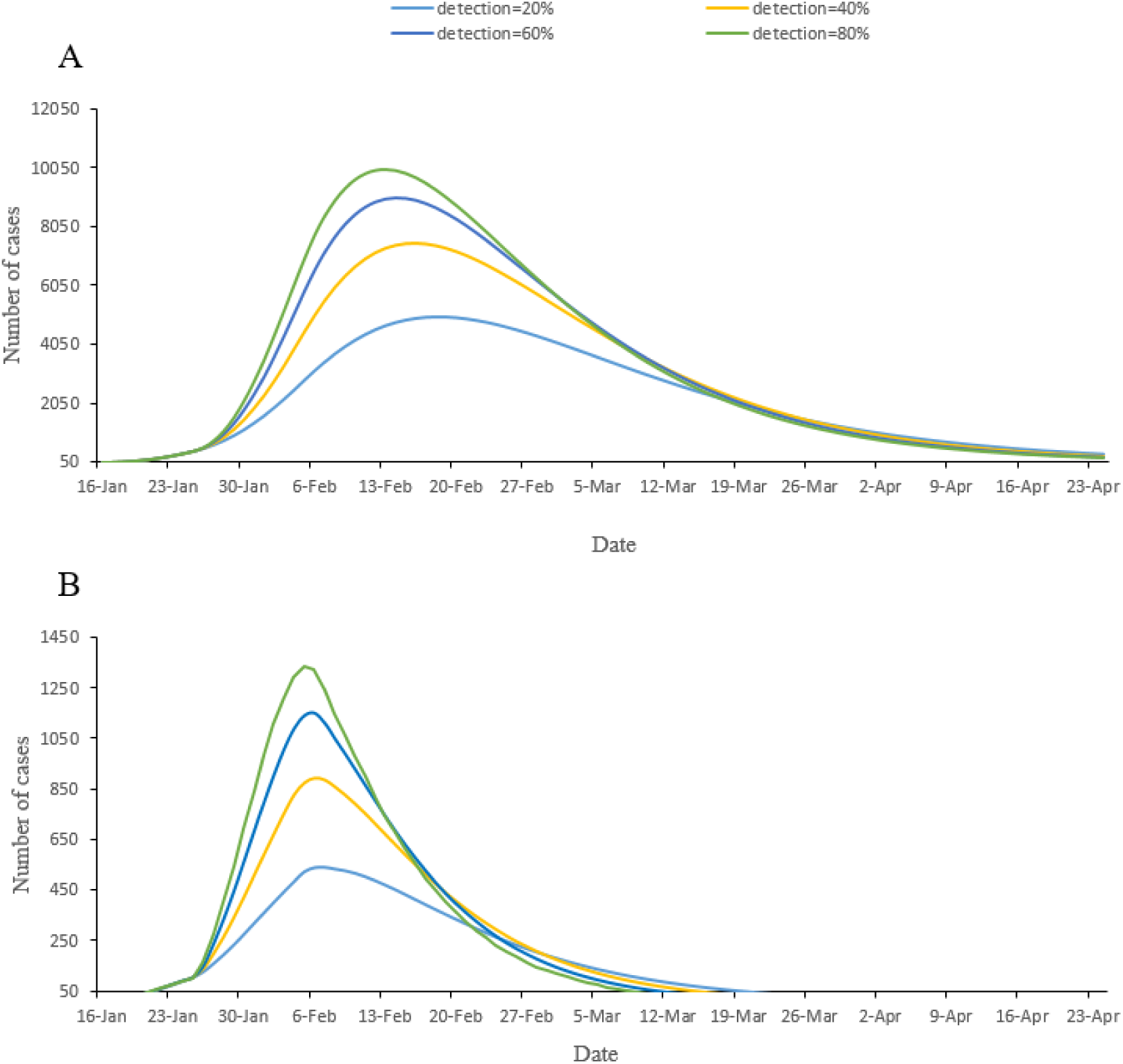
Prediction of new confirmed cases and total cases per day in hospitals, under different detection rate. A and B are used to predict the influence of different detection rates on the number of real time inpatients and the number of new confirmed cases per day, using the Wuhan Model.

**Suppl. Figure 5.**
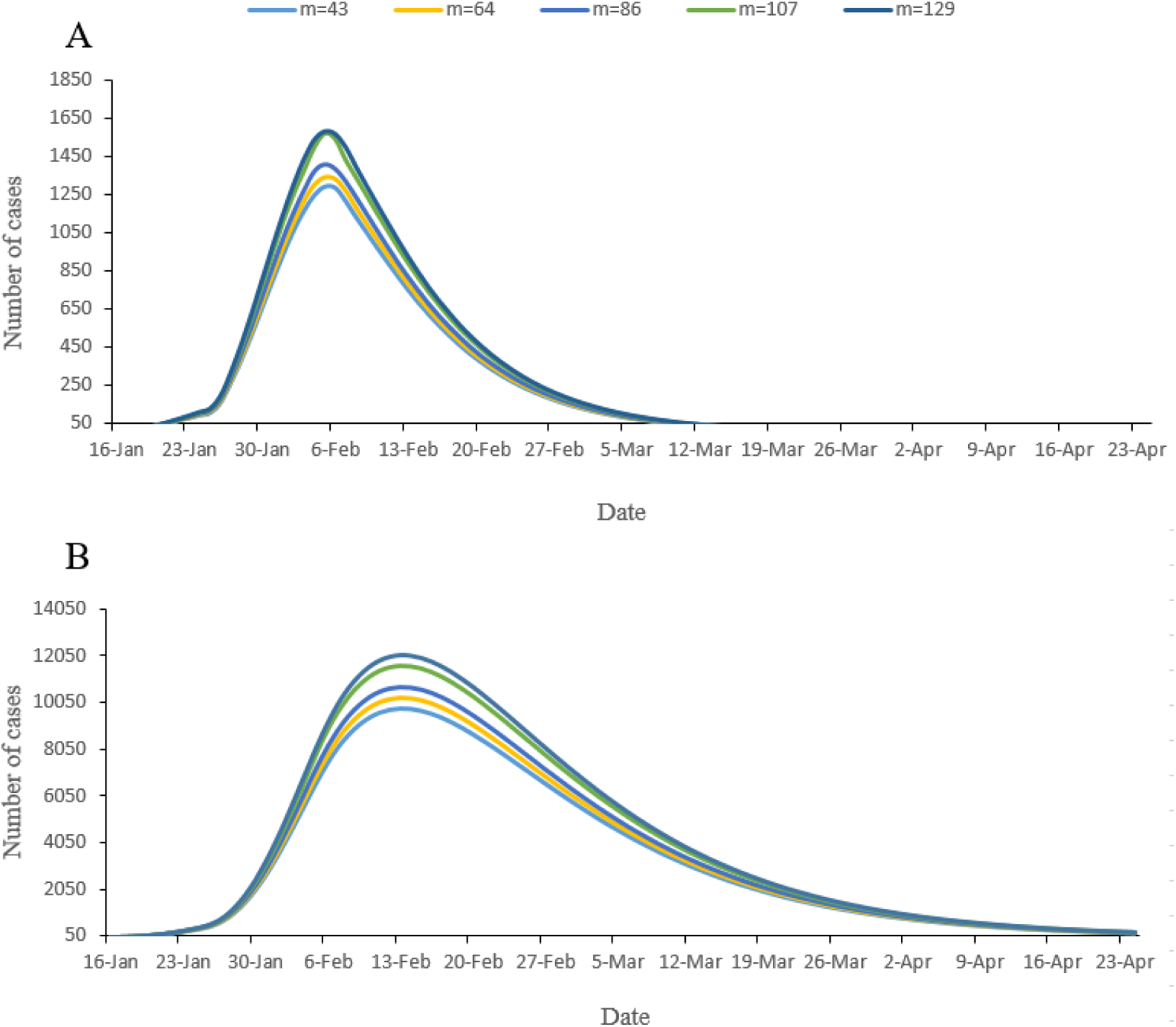
Epidemic forecasts for Wuhan under different scenarios of the number of infections in the Huanan (Southern China) Seafood Wholesale Market. The dynamic model for analyzing different situations of m, where m represents the number of confirmed cases who have been to Hainan seafood market. A and B show the prediction of the impact of the COVID-19 virus on the number of real time inpatients and newly confirmed cases per day, respectively. Under the stable outflow population, the number of newly confirmed cases per day as well as the cumulative confirmed cases have an upward trend, with an increase in the number of initial infections.

**Suppl. Figure 6.**
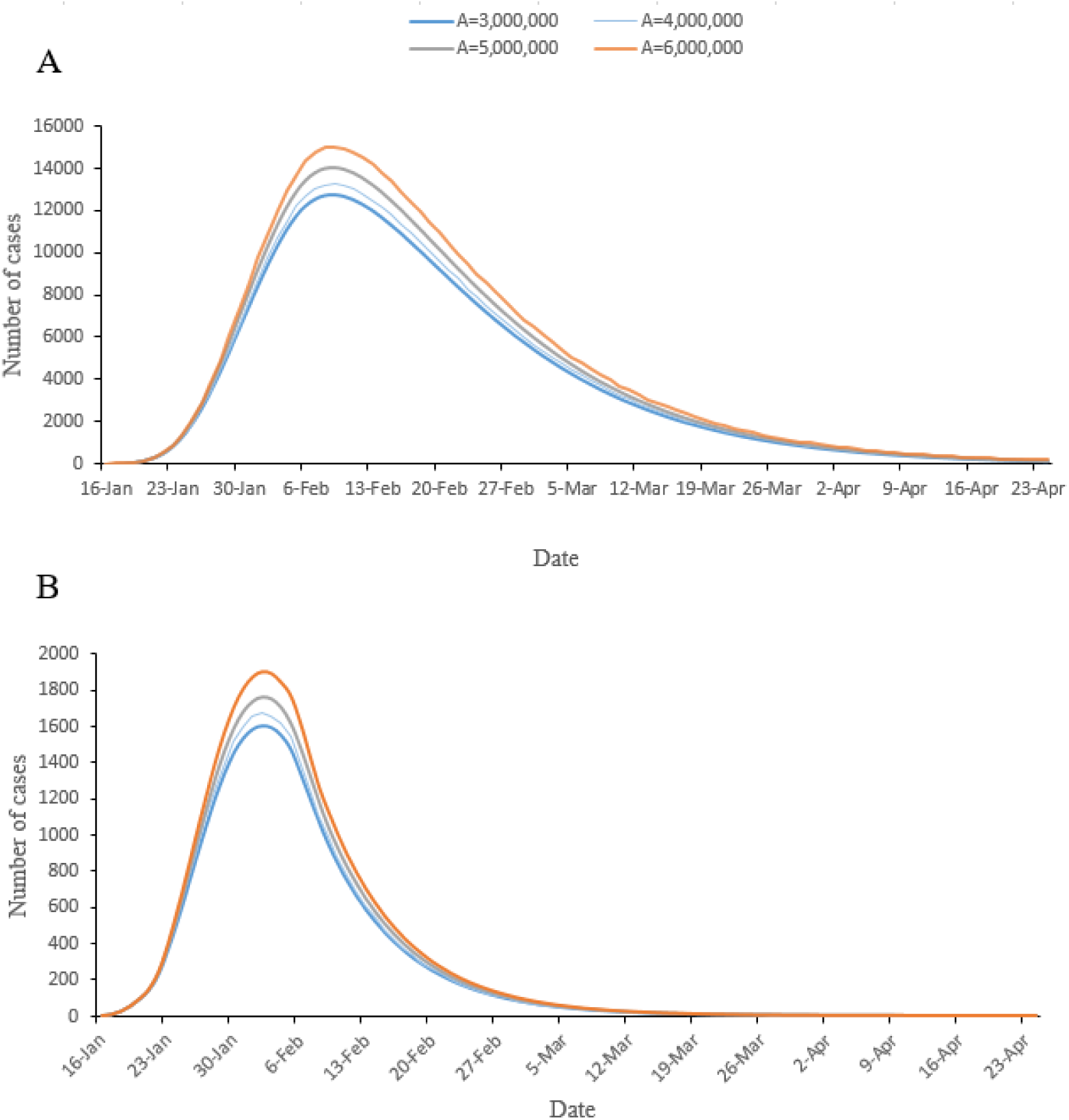
Epidemic forecasts for China (except Wuhan) under different scenarios of inflow population. We used a dynamic model to analyze various situations of **A**, where **A** represents the total population flowing from Wuhan to all parts of the country during the eight days from 16 January to 23 January 2020. We used public data to calculate the proportion of the population outflow from Wuhan, from 16 to 23January, and then allocated the proportion of daily population inflow from Wuhan to all parts of the country according to this proportion. This supplementary figure shows changes in the number of newly confirmed patients and the cumulative number of inpatients per day, assuming different levels of daily outflow population, which respectively represent the inflow population of 3,000,000, 4,000,000, 5,000,000 and 6,000,000.

**Suppl. Figure 7.**
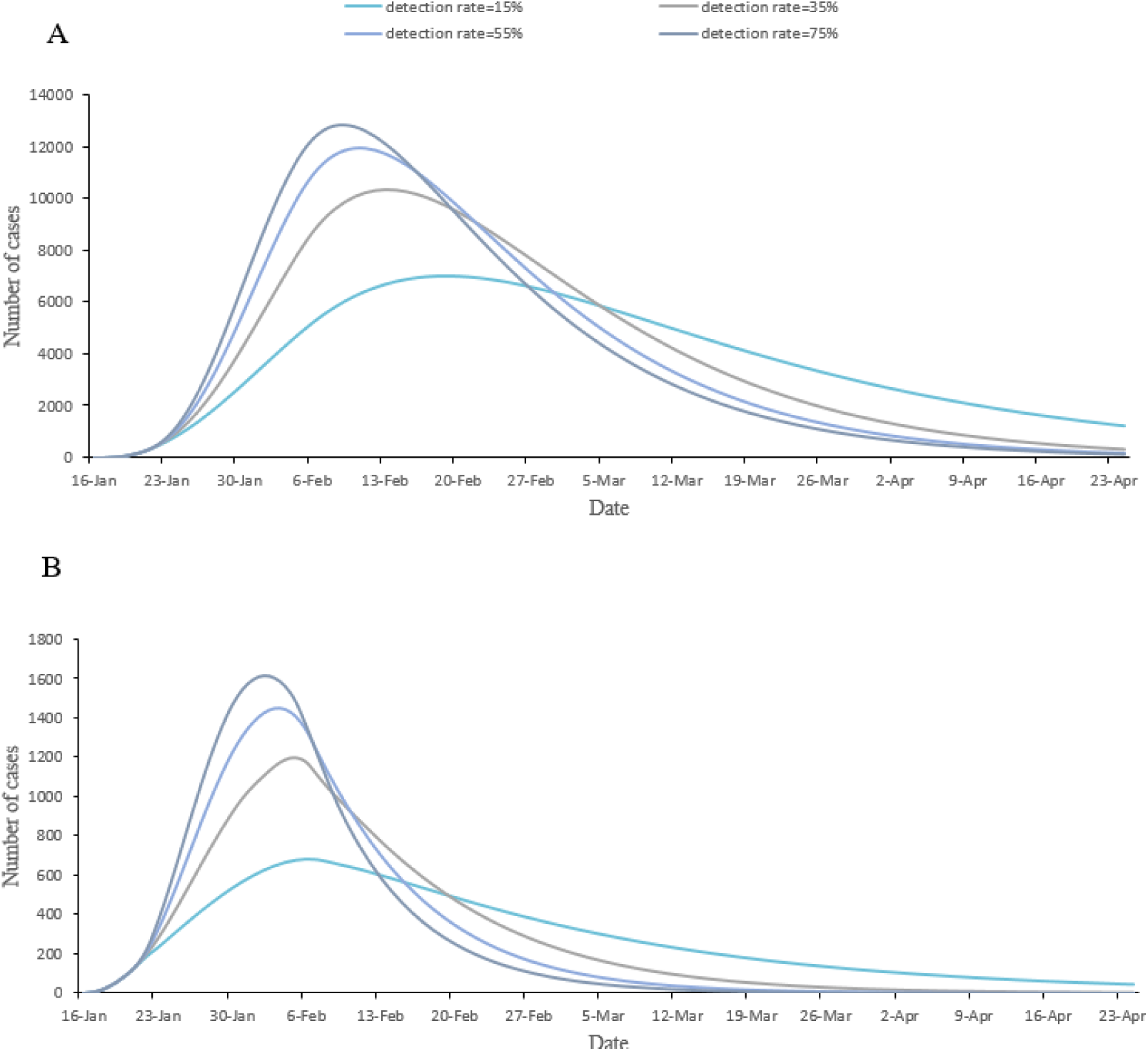
Epidemic forecasts for China (except Wuhan) under different scenarios of detection rate. A and B are used to predict the influence of different detection rates on the number of real time inpatients and the number of new confirmed cases per day, using the National Model for the whole country (except Wuhan).

## References

1. World Health Organization. Novel Coronavirus—China, Disease outbreak news : Update.https://www.who.int/csr/don/12-january-2020-novel-coronavirus-china/en/ [Accessed 28 Jan 2020].

2. Wuhan municipal health commission. http://wjw.wh.gov.cn/front/web/showDetail/2020011109035 [Accessed 28 Jan 2020].

3. Guan W-j, Ni Z-y, Hu Y, Liang W-h, Ou C-q, He J-x, et al. Clinical characteristics of 2019 novel coronavirus infection in China. medRxiv. 2020:2020.02.06.20020974.

4. Chen N, Zhou M, Dong X, Qu J, Gong F, Han Y, et al. Epidemiological and clinical characteristics of 99 cases of 2019 novel coronavirus pneumonia in Wuhan, China: a descriptive study. Lancet (London, England). 2020. Epub 2020/02/03.

5. Xinhua News. http://www.nhc.gov.cn/xcs/s3574/202002/5476aaf6e16e4e79abe7d12a317b59fc.shtml. [Accessed 28 Jan 2020].

6. World Health Organization. Novel Coronavirus (2019-nCoV) situation reports. https://www.who.int/docs/default-source/coronaviruse/situation-reports/20200206-sitrep-17-ncov.pdf?sfvrsn=17f0dca_2 [Accessed 28 Jan 2020].

7. Rock K, Brand S, Moir J, Keeling MJ. Dynamics of infectious diseases. Reports on progress in physics Physical Society (Great Britain). 2014;77(2):026602. Epub 2014/01/22.

8. National Health Commission of the People’s Republic of China. http://www.nhc.gov.cn/xcs/xxgzbd/gzbd_index.shtml [Accessed 28 Jan 2020].

9. Health Commission of Hubei Province http://wjw.hubei.gov.cn/bmdt/ztzl/fkxxgzbdgrfyyq/ [Accessed 28 Jan 2020].

10. Baidu migration map. http://qianxi.baidu.com. [Accessed 28 Jan 2020].

11. Aaron A. King. Introduction to model parameter estimation (October 30, 2017). Available at: http://creativecommons.org/licenses/by-nc/3.0/.

12. Wu JT, Leung K, Leung GM. Nowcasting and forecasting the potential domestic and international spread of the 2019-nCoV outbreak originating in Wuhan, China: a modelling study. Lancet (London, England). 2020. Epub 2020/02/06.

13. Li Q, Guan X, Wu P, Wang X, Zhou L, Tong Y, et al. Early Transmission Dynamics in Wuhan, China, of Novel Coronavirus-Infected Pneumonia. The New England journal of medicine. 2020. Epub 2020/01/30.

14. Breban R, Riou J, Fontanet A. Interhuman transmissibility of Middle East respiratory syndrome coronavirus: estimation of pandemic risk. Lancet (London, England). 2013;382(9893):694-9. Epub 2013/07/09.

15. Cheng VCC, Wong SC, To KKW, Ho PL, Yuen KY. Preparedness and proactive infection control measures against the emerging Wuhan coronavirus pneumonia in China. The Journal of hospital infection. 2020. Epub 2020/01/22.

16. Jin YH, Cai L, Cheng ZS, Cheng H, Deng T, Fan YP, et al. A rapid advice guideline for the diagnosis and treatment of 2019 novel coronavirus (2019-nCoV) infected pneumonia (standard version). Military Medical Research. 2020;7(1):4. Epub 2020/02/08.

17. Wang D, Hu B, Hu C, Zhu F, Liu X, Zhang J, et al. Clinical Characteristics of 138 Hospitalized Patients With 2019 Novel Coronavirus-Infected Pneumonia in Wuhan, China. Jama. 2020. Epub 2020/02/08.

18. Dennis Lo YM, Chiu RWK. Racing towards the development of diagnostics for a novel coronavirus (2019-nCoV). Clinical chemistry. 2020. Epub 2020/02/08.

19. The L. Emerging understandings of 2019-nCoV. Lancet (London, England). 2020;395(10221):311. Epub 2020/01/28.

